# A Multicenter Study of the Electrical Characteristics and Short-Term Outcomes of the Aveir VR Leadless Pacemaker

**DOI:** 10.64898/2026.03.06.26347827

**Authors:** Jian-du Yang, Ruo-gu Li, Xing-bin Liu, Xiao-lin Xue, Jiang-hua Zhang, Yong-mei Hu, Bao-jian Zhang, Lin Tong, Hui Luo, Min Shen, Zhong-li Chen, Xiahenazi Aiyasiding, Meng-xing Cai, Xiaojian Chi, Yan Dai, Baopeng Tang, Ke-ping Chen

**Affiliations:** State Key Laboratory of Cardiovascular Disease, Arrhythmia Center, Fuwai Hospital, National Center for Cardiovascular Diseases, Chinese Academy of Medical Sciences and Peking Union Medical College, Beijing, China; Department of Cardiology, Shanghai Chest Hospital, Shanghai Jiao Tong University, 241 West Huaihai Road, Shanghai, China; Department of Cardiology, West China Hospital of Sichuan University, No.37 Guoxue Alley, Chengdu, China; Department of Cardiology, the First Affiliated Hospital, Xi’an Jiaotong University, Xi’an, China; Department of Cardiology, First Affiliated Hospital of Xinjiang Medical University, 137 Liyushan South Road, Urumqi, Xinjiang, China; Department of Cardiology, The Second People’s Hospital of Chengdu, Chengdu, Sichuan, China; State Key Laboratory of Pathogenesis, Prevention and Treatment of High Incidence Diseases in Central Asia, Urumqi, China; Department 5 of Cardiology, Affiliated Hospital of Traditional Chinese Medicine of Xinjiang Medical University, Urumqi, China; Department of Cardiology, The Third People’s Hospital of Chengdu, Affiliated Hospital of Southwest Jiaotong University, Chengdu, China; Department of Cardiology, Changsha First Hospital, Changsha, China; Department of Cardiology, Xijing Hospital, Fourth Military Medical University, 127 West Changle Road, Xi’an, Shaanxi, China

**Keywords:** Aveir leadless pacemaker, Current of injury, Commanded Electrogram, Intracardiac Electrogram Duration, Short-term outcome

## Abstract

**Background:** The Aveir leadless pacemaker employs an active fixation method, enabling real-time monitoring of electrical parameters during implantation. However, comprehensive studies regarding the electrical parameters during this procedure are rare.

**Objective:** This study aims to analyze the electrical characteristics to further guide the implantation strategy and improve device stability and safety.

**Methods:** This multi-center retrospective study enrolled 119 patients (mean age 70.18 years; 59.58% female) who received the Aveir VR leadless pacemaker from November 2024 to May 2025 across ten centers in China. Intraprocedural variations in commanded electrogram (CEGM), current of injury (COI), impedance, pacing threshold, and sensing parameters were meticulously documented.

**Results:** CEGM mapping demonstrated various morphologies (R, RS, QR, QRS, and QS) aiding localization. During fixation, 58.82% of patients exhibited an increased COI from mapping to 0.5 turns, which was associated with reduced short-term pacing thresholds. From 0.5 to 1 turn, 52.94% showed further COI increases. ROC analysis revealed that an impedance increase has predictive value for short-term pacing thresholds, with an AUC of 0.634 and a cut-off value of 230 Ω (sensitivity 0.622, specificity 0.41). Lead stability showed a moderate correlation with impedance increase (ρ=0.44, P<0.001), while the correlation with COI was weak.

**Conclusion:** During Aveir implantation, CEGM variations guide site localization. Initial COI increases (0-0.5 turns) are linked to optimal short-term thresholds. Monitoring impedance increase is vital, as a threshold of 230 Ω serves as a key indicator of device stability and fixation quality.

## Introduction

Since the first pacemaker implantation in 1958^1^, pacing technology for the management of bradyarrhythmias has advanced rapidly, with nearly one million devices now being implanted annually worldwide^2^. Despite ongoing technological innovations, transvenous pacing systems continue to pose risks of peri-procedural and post-procedural complications, including pneumothorax, cardiac perforation, and venous obstruction. Leadless pacemakers present an ideal alternative to conventional transvenous pacing systems^3^.

As the first commercially available leadless pacemaker, Micra has created more opportunities for many patients unsuitable for transvenous pacemaker implantation^4^. However, due to its passive fixation design, it does not permit the assessment of electrical parameters prior to deployment. Electrical parameters can only be evaluated after the device is fully released, which may increase the likelihood of repeated deployments and repositioning.

The Aveir pacemaker can alter this situation by employing an active fixation mechanism that enables commanded electrogram (CEGM) and electrical parameter testing—including changes to COI, impedance, sensing, and pacing thresholds—both before and during fixation^5^. Theoretically, it could provide the operator with more data for decision-making and better align with established monitoring practices for active-fixation electrodes^6^. However, it is noteworthy that, following the publication of the Aveir DR i2i Clinical Trial, research on these intraoperative electrical parameters in multicenter studies has remained relatively sparse. Indeed, real-world clinical evidence in this specific area remains limited. Therefore, this study aims to investigate the clinical significance of electrical characteristics in guiding Aveir implantation across a large-scale cohort at multiple centers.

## Methods

### Study Design and Population Characteristics

This investigator-initiated, non-randomized, retrospective multi-center cohort study enrolled consecutive patients undergoing Aveir VR leadless pacemaker implantation at ten centers across China. Patients lacking intraoperative CEGM and electrical parameters were excluded. All patients provided written informed consent prior to the procedure. The research protocol employed in this study was reviewed and approved by the institutional review board of each participating hospital.

### Implantation Procedures

Serial electrical parameters and CEGM were collected during key procedural phases: mapping, fixation, tether mode, and post-release, with a final assessment on 24 hours after the procedure. All procedures were performed via a standard right femoral venous access approach with protocol-guided anticoagulation management. Device fixation was achieved through slow clockwise rotation, with electrical data — including CEGM, impedance, and pacing capture threshold (PCT)—recorded at 0.5, 1, 1.25, and 1.5 turns. In tether mode, a deflection stress test was conducted to confirm stability. The decision to reposition was at the operator’s discretion, informed by a comprehensive assessment of all parameters. A PCT greater than 2V@0.4ms prompted re-measurement after 3 minutes, while a persistent post-deployment threshold greater than 1.5V@0.4ms warranted consideration for device retrieval. The access site was closed using a figure-of-eight suture or manual compression.

### Electrophysiological Measures

The COI was manually measured based on two parameters: (1) the intracardiac electrogram duration (IED), expressed by milliseconds (ms), and (2) the amplitude of ST-segment elevation, measured by millivolts (mV), compared to the baseline value^7^.

Definition of COI increase: An increase is defined as a rise in either the COI amplitude or duration, or both. If one parameter decreases while the other increases, it still counts as an overall increase (Figure 1 A1-A2).

**Figure 1.**
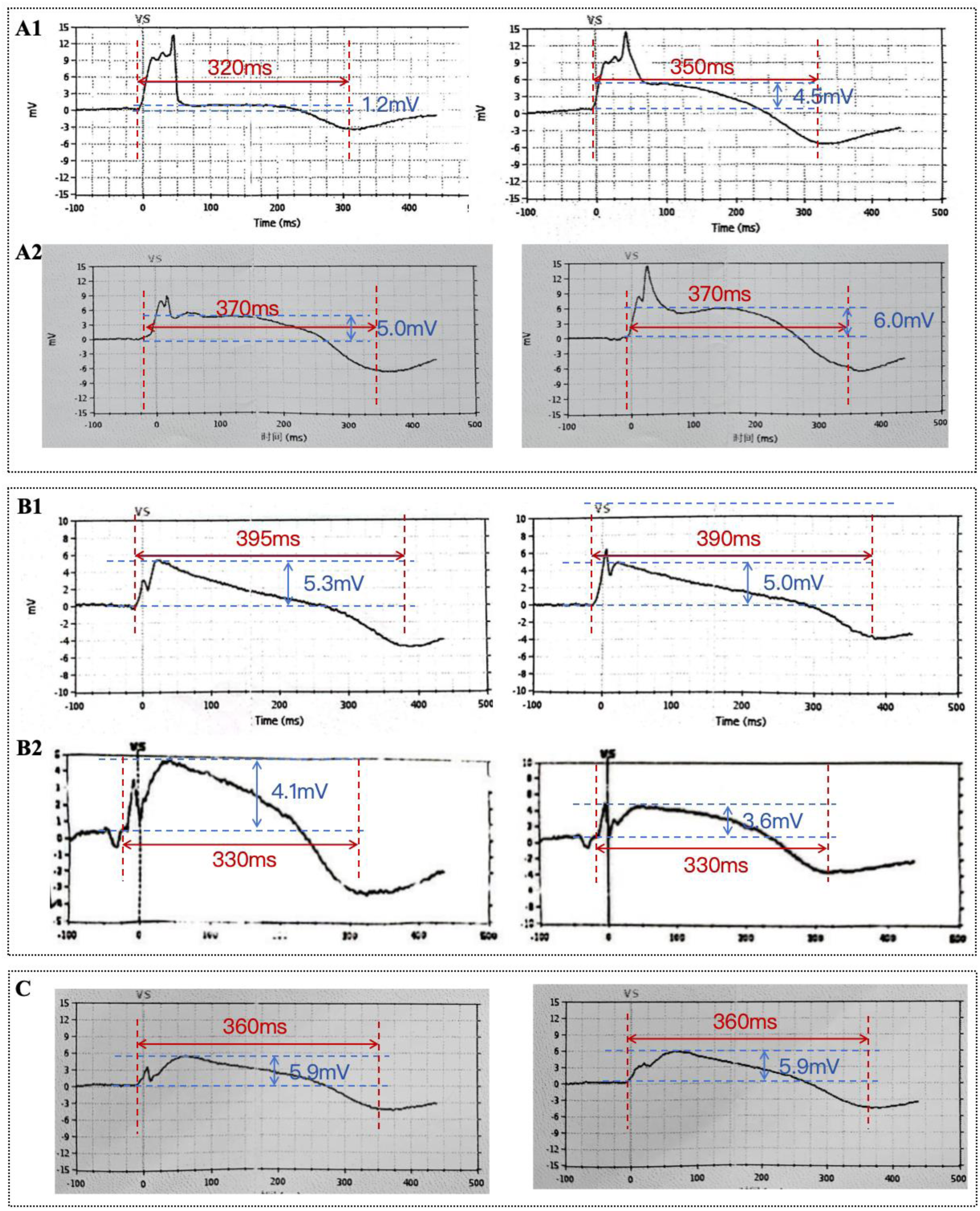
Panel A1 illustrates CEGMs in which both the COI amplitude and the IED (intracardiac electrogram duration) increase during the Aveir implantation. Panel A2 demonstrates an increase in COI amplitude while the IED remains stable. Panel B1 depicts an increase in COI amplitude with a shortening of the IED. Panel B2 shows a decrease in COI amplitude only. Panel C indicates that the COI remains stable.

Definition of COI decrease: A decrease is defined as a reduction in both the COI amplitude and IED, or if one parameter remains unchanged while the other decreases (Figure 1 B1-B2).

The definition of a stable COI indicates that no change is observed when both the amplitude and IED remain consistent relative to the baseline (Figure 1C).

### Statistical Analysis

Categorical variables were presented as frequencies and percentages and were compared using Pearson’s χ test. If any cell count was 5 or less, Fisher’s exact test was used instead. Continuous variables were tested for normality. The Shapiro-Wilk test was used to assess whether the data followed a normal distribution. Normally distributed data were expressed as the mean ± standard deviation (SD) and compared with a Student’s t-test. Non-normally distributed data were presented as the median (interquartile range) and compared using the Mann–Whitney U test. The correlation between pacing threshold and implant characteristics was assessed using Spearman’s correlation test. All statistical tests were two-tailed, and a P value < 0.05 was considered statistically significant. Receiver operating characteristic (ROC) analysis was performed to determine the predictive power of the variables. All analyses were conducted using SPSS version 29.0.

## Results

### Study population and baseline characteristics

Between November 2024 and May 2025, 119 patients underwent implantation of a single-chamber Aveir VR system. The indications for pacemaker implantation were as follows: 55 patients received the device due to sick sinus syndrome, and 64 patients due to atrioventricular block (AVB). Among these, 10 patients were diagnosed with dual nodal dysfunction. The baseline characteristics of the patients are presented in Table 1.

**Table 1.**
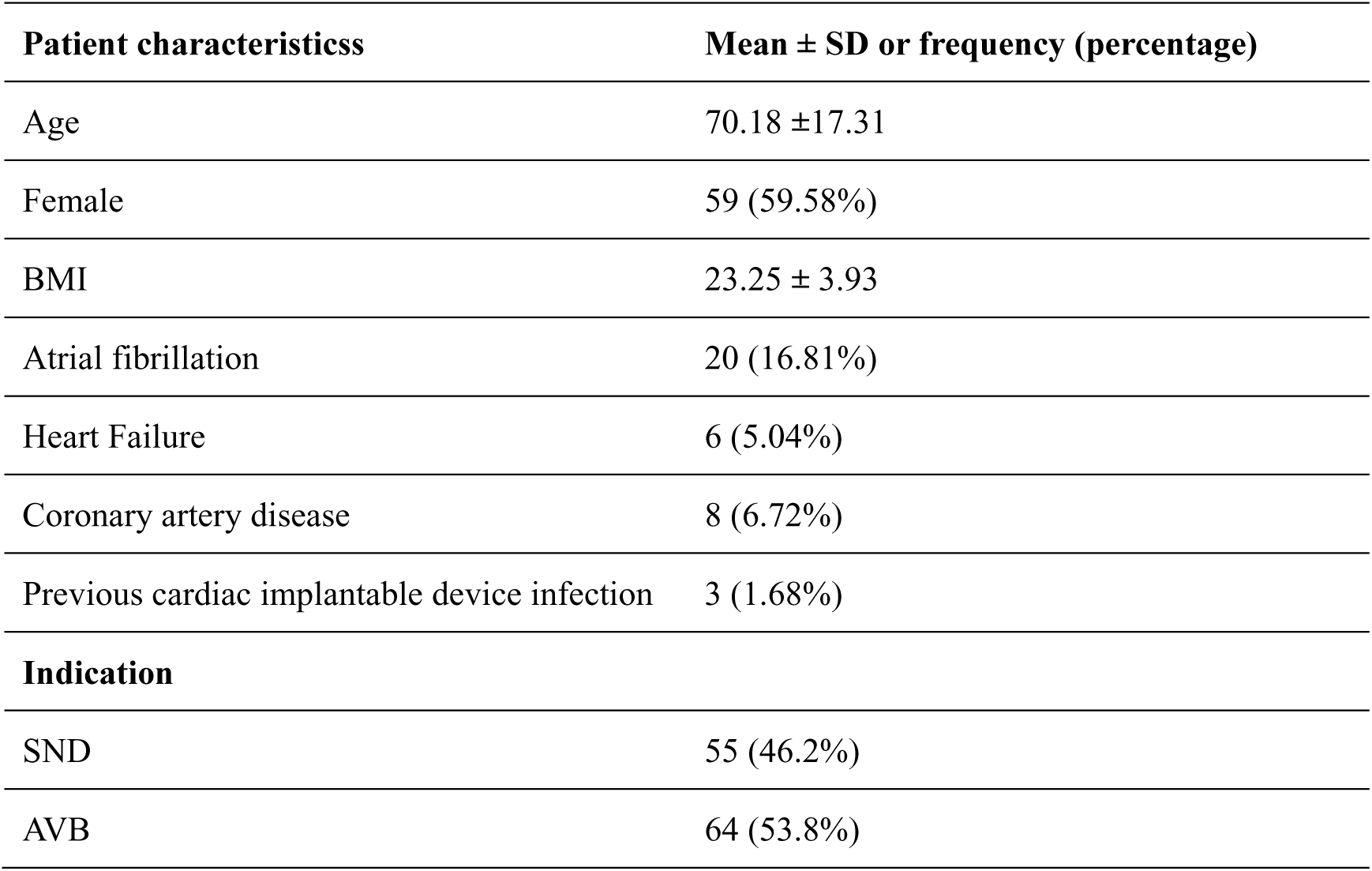
Characteristics of the Aveir VR Patient Cohort.

| Patient characteristicss | Mean ± SD or frequency (percentage) |
| --- | --- |
| Age | 70.18 ± 17.31 |
| Female | 59 (59.58%) |
| BMI | 23.25 ± 3.93 |
| Atrial fibrillation | 20 (16.81%) |
| Heart Failure | 6 (5.04%) |
| Coronary artery disease | 8 (6.72%) |
| Previous cardiac implantable device infection | 3 (1.68%) |
| **Indication** |  |
| SND | 55 (46.2%) |
| AVB | 64 (53.8%) |

### General Procedure Characteristics

The median implantation and fluoroscopy times were 45.0 minutes (interquartile range [IQR]: 33.0–55.0) and 10 minutes (IQR: 6–12.5), respectively. The overall implant success rate was 100%. Thirteen patients (10.92%) required device repositioning. At the conclusion of the procedure, the median sensing, impedance, and pacing threshold values were 7.2 mV (IQR: 5.1–9.9), 700 Ω (IQR: 515–907.5 Ω), and 0.5 mV (IQR: 0.5–0.75), respectively, with a pulse width of 0.4 ms.

Of all patients, 99 (83.19%) received the Aveir device positioned at the inferior septal region of the right ventricle, 9 patients (7.56%) at the mid-septal area of the right ventricle, 4 patients (3.36%) at the apex of the right ventricle, and 7 patients (5.88%) at the free wall of the right ventricle. No significant periprocedural complications were observed.

### CEGM Morphology Characteristics

In this study, statistical analysis was conducted on CEGMs across all patients (mapping was attempted 136 times, with 150 fixation instances). During the mapping process, the CEGM morphology was identified as RS in 69 patients (57.98%), R in 42 patients (35.29%), QR in 6 patients (5.04%), QRS in 1 patient (0.84%), and QS in 1 patient (0.84%) (Figure 2). The morphology of CEGMs varies according to the implantation site. The low septum, mid-septum, and free wall predominantly exhibit an RS morphology, accounting for 57.58%, 66.67%, and 71.43% of cases, respectively, whereas the apex mainly displays an R-wave morphology, representing 75%. The Chi-square test indicated a statistically significant difference between the groups, with a P-value of <0.0001 (Table 2).

**Figure 2.**
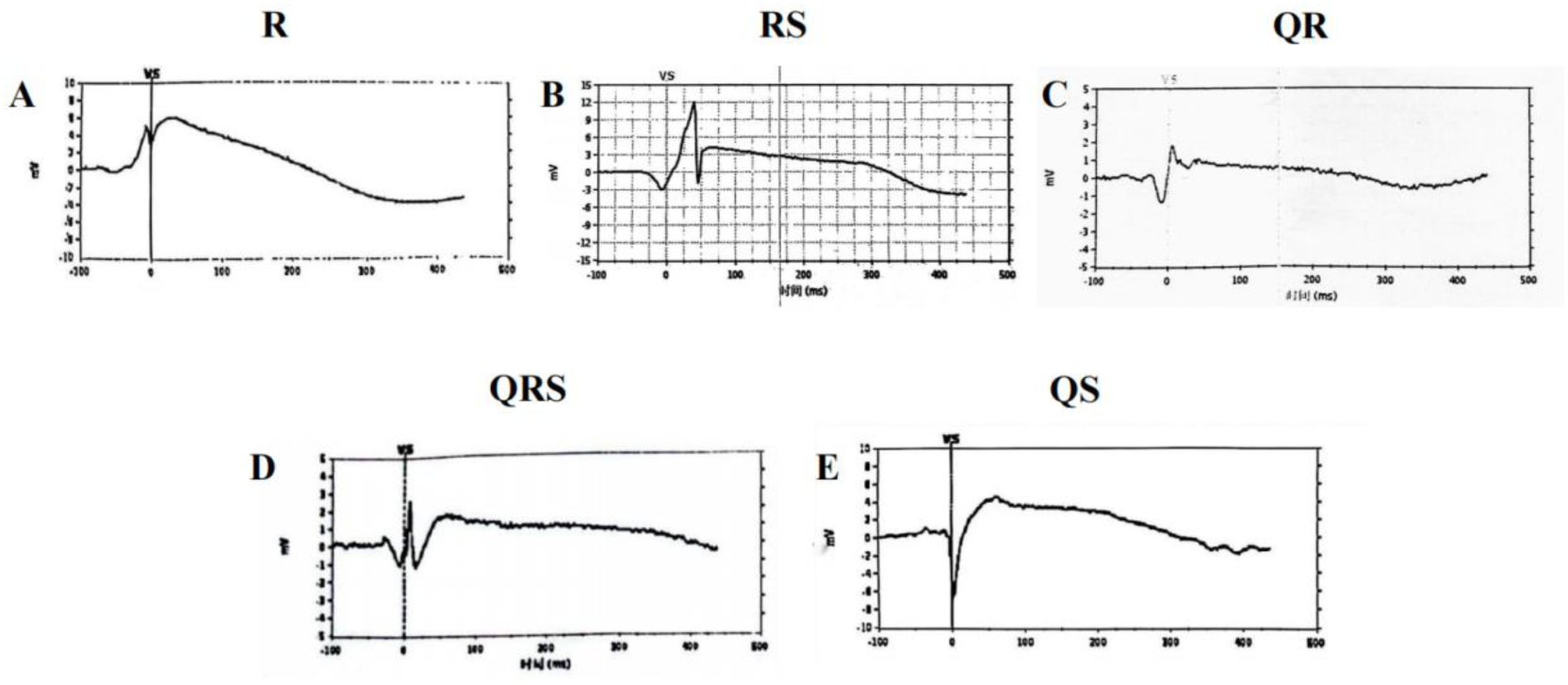
Panel A shows the R morphology of the CEGM, Panel B shows the RS morphology, Panel C shows the QR morphology, Panel D shows the QRS morphology, and Panel E shows the QS morphology.

**Table 2.** CEGM morphologies and Aveir pacing parameters recorded at various Aveir implantation locations.

|  | Low-septal | Mid-septal | Apex | Free-wall | P-value |
| --- | --- | --- | --- | --- | --- |
| CEGM Morphology |  |  |  |  |  |
| QR | 5.05% | 0.00% | 0.00% | 14.29% | <0.0001 |
| QRS | 0.00% | 11.11% | 0.00% | 0.00% |  |
| QS | 0.00% | 11.11% | 0.00% | 0.00% |  |
| R | 37.37% | 11.11% | 75.00% | 14.29% |  |
| RS | 57.58% | 66.67% | 25.00% | 71.43% |  |
| Pacing Parameters (implantation ) |  |  |  |  |  |
| Threshold(mV) | 0.5<br>(IQR 0.5-0.75) | 0.5<br>(IQR 0.5-0.75) | 0.5<br>(IQR 0.44-0.56) | 0.5<br>(IQR 0.5-0.75) | 0.33 |
| Sense(V) | 8.0<br>(IQR5.5-11.4) | 7.0<br>(IQR 5.1-11.9) | 6.5<br>(IQR 4.55-8.83) | 5.0<br>(IQR 4.0-12.6) | 0.53 |
| Impedance(Ω) | 690<br>(IQR 520-920) | 690<br>(IQR 680-820) | 950<br>(722.5-1157.5) | 670<br>(IQR 565-970) | 0.24 |
| Pacing Parameters (24h) |  |  |  |  |  |
| Threshold(mV) | 0.623<br>(IQR 0.5-0.75) | 0.75<br>(IQR 0.5-0.75) | 0.75<br>(IQR 0.63-0.75) | 0.63<br>(IQR 0.5-1.13) | 0.85 |
| Sense(V) | 7.7<br>(IQR 5.7-9.35) | 6.75<br>(IQR 4.75-10.45) | 9.9<br>(IQR 8.3-11.5) | 7.7<br>(IQR 6.95-8.45) | 0.52 |
| Impedance(Ω) | 670<br>(IQR 550-757) | 510<br>(IQR 440-730) | 770<br>(IQR 640-1010) | 695<br>(IQR 472-820) | 0.48 |

The pacing thresholds at the time of implantation and on the subsequent day were as follows across various QRS morphologies: RS morphology: 0.5 mV (IQR: 0.25–0.5) at implantation and 0.5 mV (IQR: 0.5–0.5) on the following day. R morphology: 0.5 mV (IQR: 0.5–0.75) at implantation and 0.75 mV (IQR: 0.5–1.0) on the subsequent day. QR morphology: 0.75 mV (IQR: 0.56–0.75) at implantation and 0.75 mV (IQR: 0.69–0.81) on the following day. QRS morphology: 0.5 mV at implantation and 0.5 mV on the following day. QS morphology: 0.5 mV at implantation and 1 mV on the following day. No statistically significant differences were observed between the various morphologies concerning pacing thresholds at the time of implantation (P=0.85) and on the following day (P=0.37). Similarly, the pacing parameters of

Aveir at different implantation sites are compared in Table 2, and no statistically significant differences were observed across all parameters.

### Relationship between COI and Pacing Threshold

The COI was documented during two phases of fixation: from mapping to 0.5 turn, and from 0.5 to 1 turn. The CEGM from 1 turn to 1.25/1.5 turns was not recorded for certain patients, thereby precluding a statistical analysis of the COI change during the final turn.

Throughout the mapping period, COI was present in all patients; however, the amplitude of COI and IED showed variability among individuals. The median amplitude of COI during mapping was 3 mV (interquartile range: 2–4.5 mV), and the mean IED was 342.85 ± 41.60 ms.

From mapping to 0.5 turn, an increase in COI was observed in 70 patients (58.82%). Among these, 55 patients (46.22%) exhibited an increase in amplitude, with a median increase of 1 mV (IQR: 1–2.5 mV), while 33 patients (27.73%) showed prolonged IED, with a median prolongation of 25 ms (IQR: 20–45 ms). A total of 18 patients (15.13%) showed increases in both the amplitude of the COI and IED duration. Conversely, 27 patients (22.69%) displayed no significant alterations in the COI. Notably, 22 patients exhibited a decrease in COI during fixation: 3 experienced reductions in both amplitude and IED, 4 showed unchanged amplitude with shortened IED, and 15 exhibited reduced amplitude with unchanged IED. The median decrease in COI amplitude was –1.25 mV (IQR: –2 to –1 mV), and the median reduction in IED duration was –30 ms (IQR: –35 to –20 ms).

During the device rotation from 0.5 to 1 turn, an increase in COI was observed in 63 patients (52.94%). Among these, 42 patients (35.29%) exhibited only an increase in amplitude, with a median increase of 1 mV (IQR: 0.55–2.0 mV). Twelve patients (10.08%) demonstrated an increase exclusively in IED, with a median increase of 25 ms (IQR: 23.75–35 ms). Nine patients (7.56%) showed increases in both amplitude and IED. Conversely, 27 patients (22.69%) exhibited no significant changes in COI during this phase. Consistent with the trend observed from mapping to 0.5-turn, a decrease in COI was documented in 29 patients (24.37%) during further fixation. Of these, 3 patients (2.52%) experienced reductions in both amplitude and duration; 6 patients (5.04%) maintained unchanged amplitude with shortened IED; and 20 patients (16.81%) demonstrated reduced amplitude with unchanged IED. The median reduction in amplitude was –1 mV (IQR: –2 to –0.5 mV), and the median decrease in IED was –25 ms (IQR: –40 to –20 ms).

In summary, during the device rotation from the mapping phase to 1 turn, the majority of patients exhibited an increase in COI. However, a small subset of patients (17 patients, 14.29%) demonstrated no change or a decrease. Among these, 7 patients (5.88%) showed no significant change in COI, whereas 10 patients (8.40%) experienced a reduction (Supplementary Figure 1).

The variations in COI fluctuate throughout each screwing phase and can exhibit multiple patterns, as illustrated in Supplementary Figure 1. The correlation analysis with the threshold during Aveir implantation and the next-day thresholds is as follows: patients exhibiting a decrease in COI to 0.5 turn during mapping had a significantly higher short-term (24 hours post-procedure) threshold (P = 0.02) compared to those with increased or stable COI, although no statistically significant difference was observed in the immediate post-procedure threshold (P = 0.054). Conversely, changes in COI during the 0.5–1 turn did not demonstrate any significant correlation with either the immediate (P = 0.45) or the next-day threshold (P = 0.82) (Figure 3).

**Figure 3.**
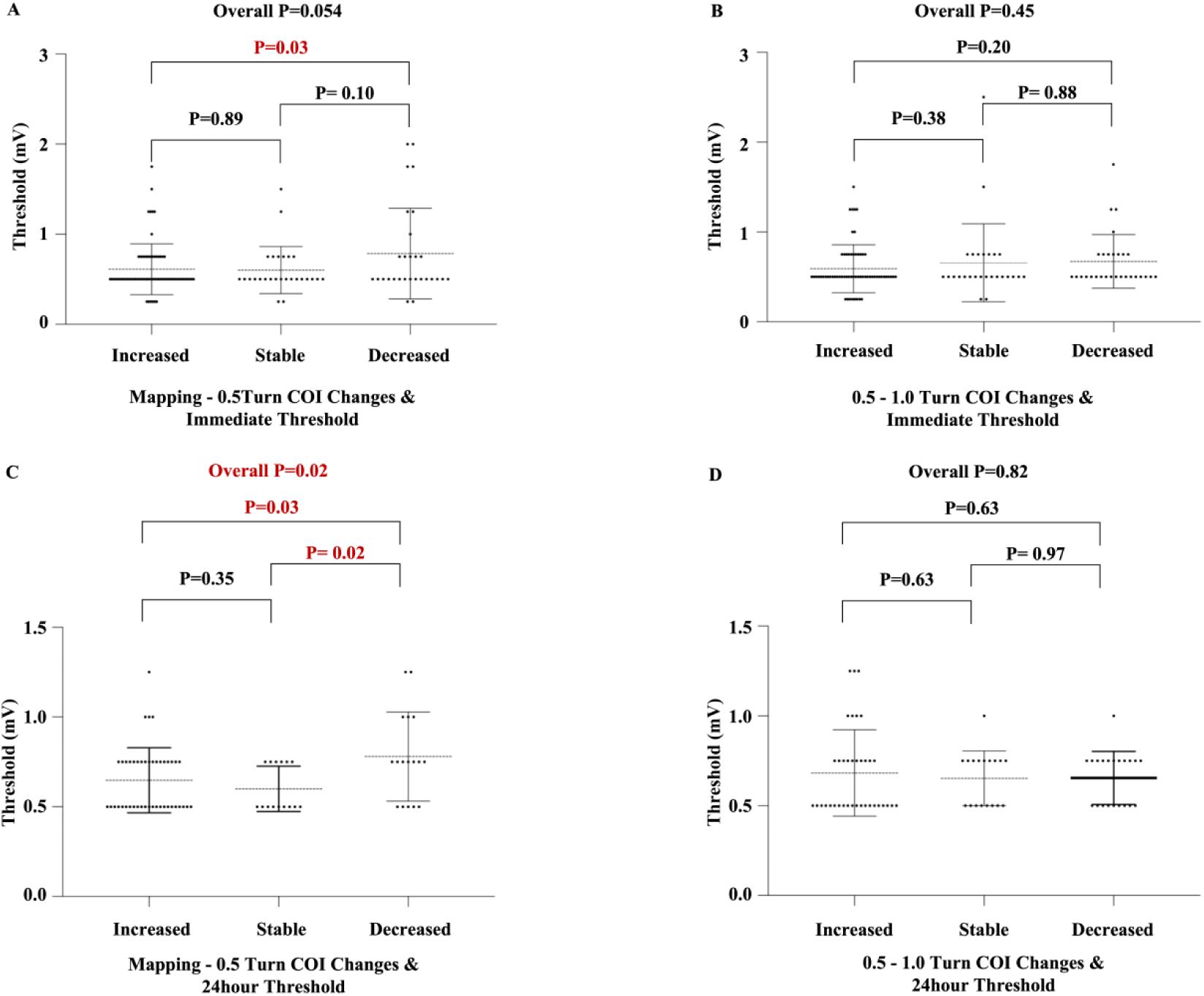
Panel A illustrates the comparison of immediate thresholds among various COI change groups from mapping to 0.5 turns. Panel B displays the comparison of immediate thresholds among different COI change groups from 0.5 to 1 turn. Panel C shows the comparison of 24-hour post-implantation thresholds among different COI change groups from mapping to 0.5 turns. Panel D presents the comparison of 24-hour post-implantation thresholds among various COI change groups from 0.5 to 1 turn.

### The Relationship Between Impedance and Pacing Threshold

During the acute phase of the implantation procedure, impedance progressively increases as the Aveir is screwed in. The threshold initially increased with the screwing-in process and then gradually decreased during the observation period after release. Sensing shows a gradual increasing trend during the screwing-in process. Twenty-four hours post-implantation, a reduction in impedance was observed, the pacing threshold gradually declined, and sensing remained largely stable (Figure 4A-C).

**Figure 4.**
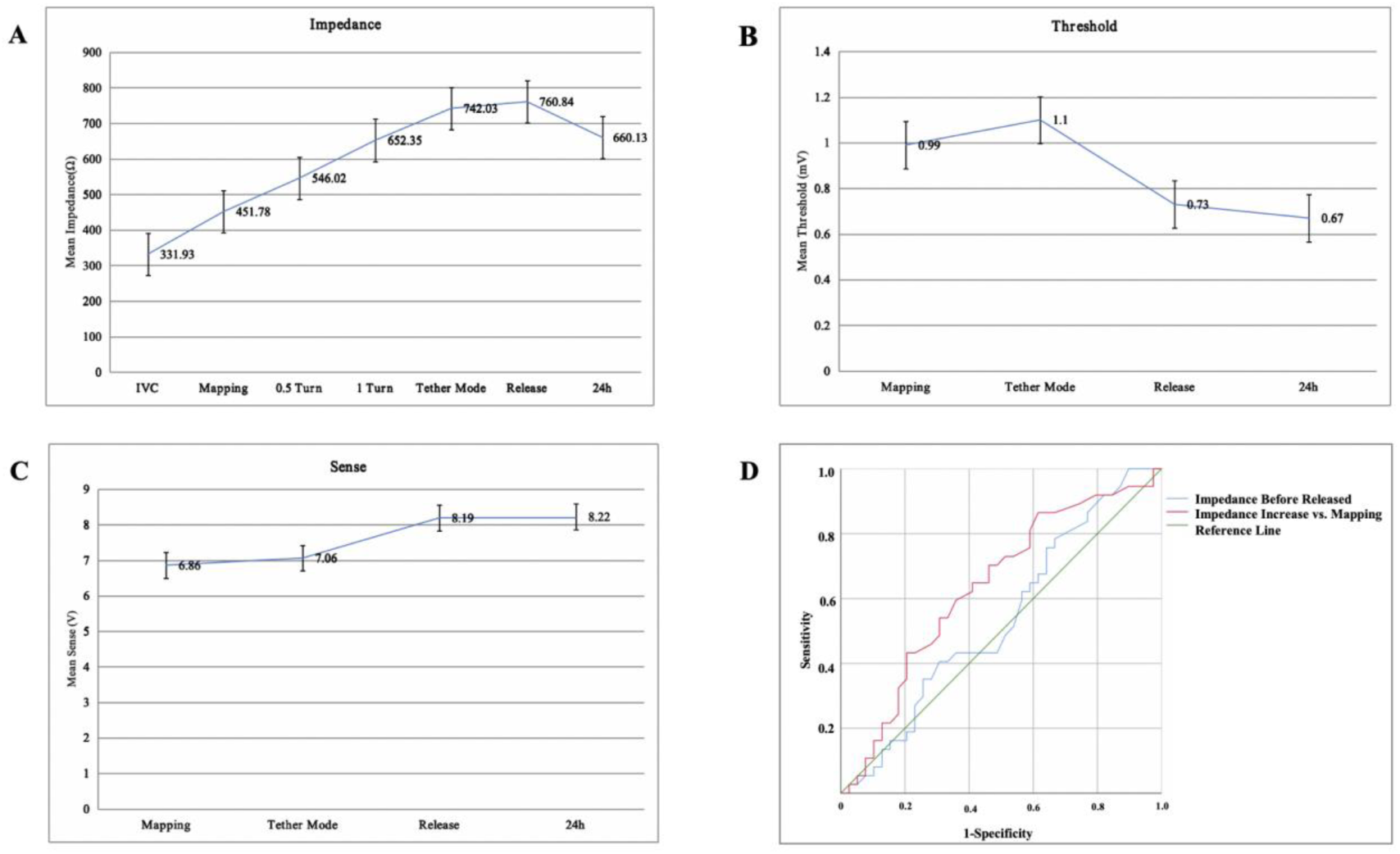
Panels A-C depict electrical measurements of Aveir at various stages of implantation and during the acute follow-up period; Panel D presents the ROC curve for intraprocedural impedance in predicting postoperative threshold.

Using ROC curve analysis (Figure 4D), the predictive value of impedance increase during device fixation and at short-term (24h post-implant) thresholds was assessed. The area under the ROC curve (AUC) for impedance increase was 0.634 (95% CI 0.406-0.667), with a cut-off value of 230 Ω (sensitivity = 0.65, specificity = 0.59). Additionally, for impedance before release, the AUC was 0.536 (95% CI 0.508-0.761), with a cut-off value of 645 Ω (sensitivity = 0.622, specificity = 0.41).

### Correlation Between Aveir Stability and Implantation Parameters

Throughout the entire implantation procedure, a total of 10 patients required repositioning due to initial implant instability. Spearman’s rank correlation analysis was conducted. Lead stability exhibited a moderate correlation with the increase in impedance relative to the IVC (R=0.44, P<0.001). Conversely, it demonstrated weak correlations with the amplitude increase of the COI during implantation (R=0.18, P=0.052) and with the prolongation of the IED (R=0.04, P=0.664).

## Discussion

The CEGM provides crucial insights during Aveir implantation, serving as a valuable tool for evaluating the optimal implantation site. While a definitive correlation between CEGM morphology and pacing threshold has not been established, it is well recognized that the excitation of the tissue in contact with the electrode tip influences the formation of CEGM. Excitation sequences differ across regions, resulting in variations in the generated CEGM, which is significant for determining the location of Aveir^8^.

According to the research by Louie Cardone-Noott et al.^9^, ventricular activation propagates along the Purkinje fibers, initially activating the left ventricular septum through the septal branches, then conducting across the septum to the right ventricular septum surface. Simultaneously, the right ventricular (RV) apex is activated first via the right bundle branch (RBB), with subsequent conduction toward the free wall. Therefore, the sequence of RV activation is as illustrated in Figure 5 (red arrows indicate the initially activated areas, and blue arrows indicate the subsequent activation areas). This difference in activation sequence results in distinct CEGM recordings across different implantation sites, allowing for precise localization. For instance, when the Aveir device is implanted in the low or mid-septum, activation originates in the LV septum and propagates toward the RV septum. As a result, the activation vector, pointing from the tip to the end of the Aveir device, produces a positive R-wave (green waveform in Figure 5A). Conversely, when the activation moves away from the Aveir tip, it creates a negative component (yellow waveform in Figure 5A). At the apex, the activation sequence becomes more complex due to multiple pathways partially canceling each other out. Still, overall activation progresses from the apex toward the base, resulting in a predominant R-wave morphology (Figure 5B). When implanted at the RV free wall, the activation pattern resembles that of septal sites, and most patients exhibit an RS morphology (shown in Figure 5C). Therefore, based on the morphology of the CEGM, physicians can identify the implantation site, which is particularly useful for patients with renal insufficiency, as it may reduce the necessity for repeated angiography.

**Figure 5.**
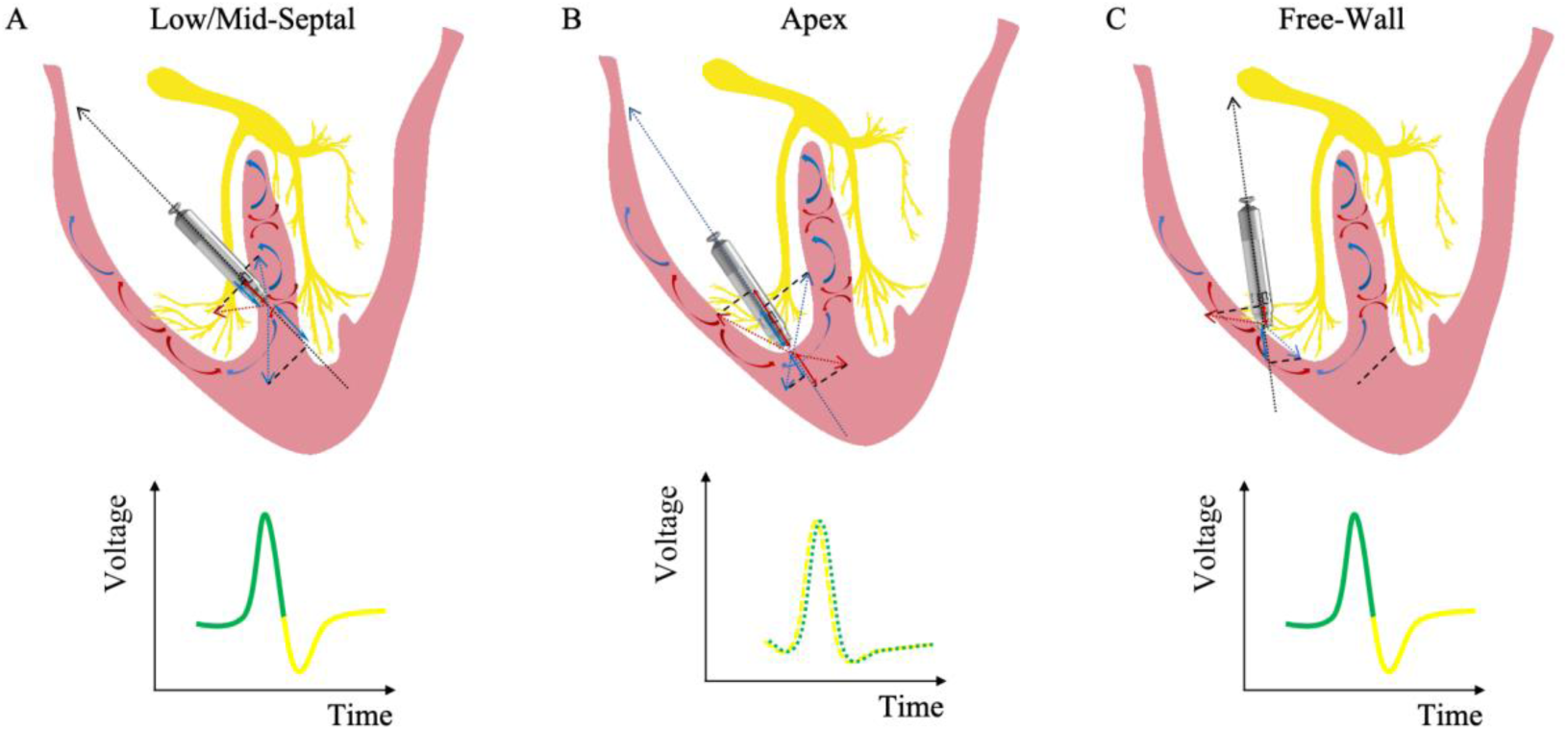
Conceptual representation of the spatial and temporal relationships for CEGM.

Based on a prospective study of 96 active-fixation leads, the research demonstrated that the presence of a COI—characterized by prolonged IED and ST-segment elevation at the time of fixation—predicts successful lead placement. Conversely, the absence of COI is associated with persistently high thresholds or acute lead dislodgement, indicating the necessity for lead repositioning. Therefore, it supports the utilization of real-time COI monitoring to assess fixation stability and inform clinical decisions during implantation^7^.

Previous studies have established that during fixation of an active lead, the COI generally increases as the lead penetrates deeper, unless it is screwed in continuously until perforation occurs, at which point the COI decreases. At this stage, the helix is positioned closer to the opposite endocardial surface, thereby supporting the hypothesis that the COI is positively correlated with the depth of helix implantation into the myocardium^10^. Similarly, this study also observed the presence of COI during Aveir implantation. However, unlike prior reports, continuous dynamic recording of the CEGM did not demonstrate a sustained increase in the injury current. Instead, some patients displayed either a stable COI or a decrease relative to the mapping phase. We hypothesize that although the Aveir is technically an active fixation device, its design features a helix primarily serving as an anchoring mechanism, whereas the cathode itself more closely resembles a passive electrode. Consequently, the recorded COI may reflect a passive tissue contact process rather than active tissue penetration. This constitutes a distinction from the design of atrial Aveir leads. We further hypothesize that the lack of further COI increase during fixation may be due to the cathode becoming lodged within myocardial trabeculations. The right ventricle contains numerous trabeculae carneae, and it is plausible that during implantation, the helix becomes embedded within a trabeculation while the tip remains confined within intertrabecular spaces. This mechanical configuration may inhibit additional tissue injury and consequently limit further increases in COI (as illustrated in Figure 6).

**Figure 6.**
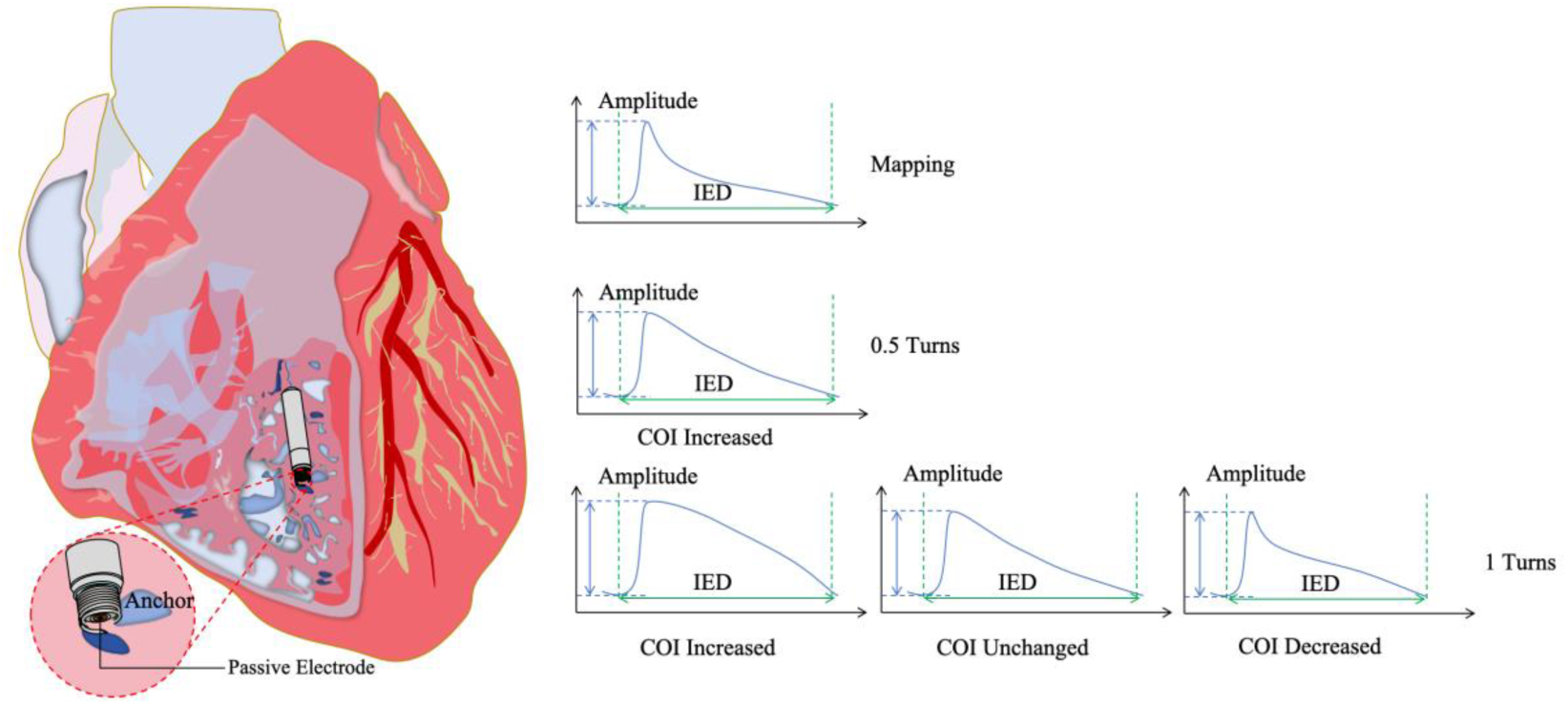
Illustrates a schematic diagram of the Aveir implantation site in the right ventricular (RV) septal region. The RV low septum often contains abundant trabeculated musculature. During helix fixation at a low septal site, the helix frequently engages with the trabeculated muscle, while the cathode tip does not penetrate deeply into the septum. Consequently, the COI often decreases or remains stable. However, due to the fixed, secure contact, increased impedance results from closer adherence between the cathode and the tissue.

This discovery does not suggest that elevated COI carries no value for short-term outcomes; conversely, our study confirmed that within the initial 0.5 turns of fixation, patients with decreased COI had significantly higher short-term threshold values than those with unchanged or increased COI. In the 0.5-1 turn fixation procedure, no significant differences in threshold were observed among groups with different COI changes. This indicates that the initial phase of fixation strongly influences the subsequent electrical parameters of the leadless pacemaker. To summarize, if no COI rise occurs in the first half-turn, immediate repositioning is advised.

Furthermore, the study established that a reduction in COI during the 0.5-1 turn process is not a decisive factor for repositioning. If COI decreases between 0.5 and 1 turn while impedance increases during this interval, additional fixation of the Aveir may be considered, with ongoing assessment of electrical parameters.

Moreover, consistent with previous studies^11^, this study confirms that impedance has considerable predictive value for determining optimal lead implantation parameters. Additionally, our findings indicate that the increase in impedance offers superior predictive efficacy compared to impedance levels alone. Given that each patient has a unique baseline impedance, the reference value depends on the individual’s baseline level. Conversely, the relative increase in impedance more precisely reflects the extent of contact between the Aveir device and the cardiac tissue. Consequently, this study concludes that utilizing the increase in impedance during the implantation procedure, in comparison to the mapping phase, as a reference indicator is more justified.

## Limitation

Due to some patients being unable to undergo follow-up, this study only investigated the perioperative and short-term electrical parameters of the leadless pacemaker; the long-term outcomes remain unclear and require further investigation.

## Conclusion

During the Aveir implantation procedure, the CEGM showcases notable variations that align with different implantation locations, offering valuable guidance for localization. Fluctuations in COI at various stages vary among individuals, with a 0-0.5 turn increase closely linked to short-term threshold values. Additionally, the degree of impedance increase during implantation is crucial, as it directly relates to both device stability and short-term thresholds post-implantation.

## Data Availability

Datasets used or analyzed during the current study are available from the corresponding author on reasonable request.

## Ethics Approval and Consent to Participate

All patients provided written informed consent. The present study was approved by the Ethics Committee of Fuwai hospital。

## Consent for publication

Not applicable.

## Clinical trial number

Not applicable.

## Authors’ contributions

Jian-du Yang collect data and write the original draft preparation; Ruo-gu Li, Xing-bin Liu,, Xiao-lin Xue, Jiang-hua Zhang revised the main manuscript text; Yong-mei Hu, Bao-jian Zhang, Lin Tong prepared the figures and tables, Hui Luo, Min Shen, Zhong-li Chen, Xiahenazi Aiyasiding, Meng-xing Cai, Xiaojian Chi, Yan Dai collected datas. All authors reviewed the manuscript.

## Acknowledgements

Not applicable.

## Abbreviations

COI: current of injury
IED: intracardiac electrogram duration
CEGM: commanded electrogram
AUC: area under curve
ROC: Receiver operating characteristic
PCT: pacing capture threshold
IQR: interquartile range
RV: right ventricular
RBB: right bundle branch;

---

**Supplementary Figure 1.**
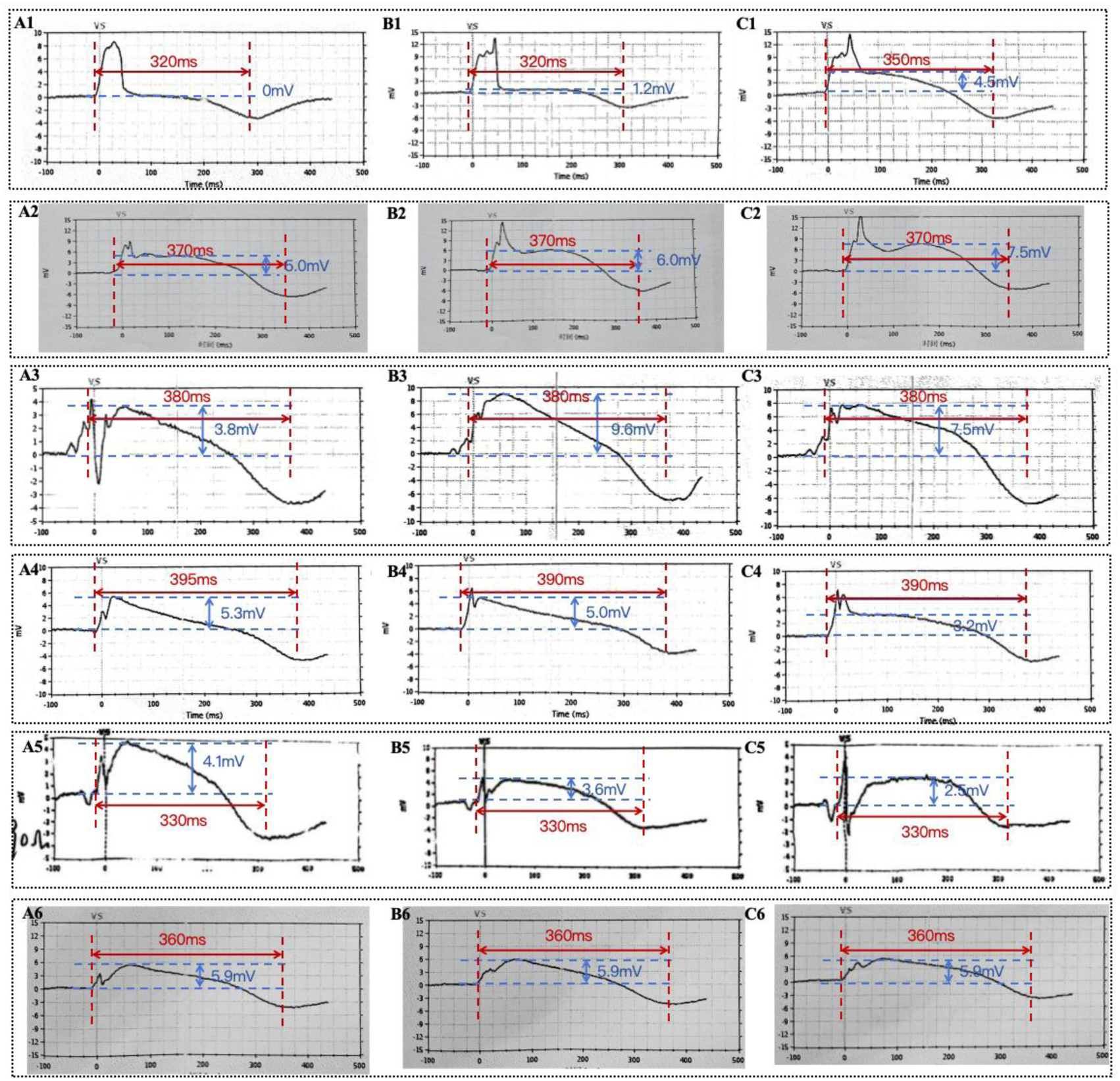
A1-C1 shows CEGMs where both the COI amplitude and the IED (Intracardiac Electrogram Duration) increase during the Aveir implantation procedure. A2-C2 illustrate increases in COI amplitude with stable IED. A3-C3 depict increases in COI amplitude whilst the IED shortens. A4-C4 demonstrates decreases in both COI and IED. A5-C5 indicate that the COI remains stable. Figures A1-6 present the CEGMs corresponding to mapping procedures; Figures B1-6 illustrate CEGMs at 0.5 turns; and Figures C1-6 display CEGMs at 1 turn.

